# Effect of outpatient cardiac rehabilitation following in-hospital stroke rehabilitation on motor function and health-related quality of life in stroke survivors

**DOI:** 10.1101/2023.11.22.23298931

**Authors:** Chiaki Yokota, Masatoshi Kamada, Kiyomasa Nakatsuka, Misa Takegami, Hiroyuki Miura, Makoto Murata, Hiroaki Nishizono, Kunihiro Nishimura, Yoichi Goto

## Abstract

**Background:** Outpatient cardiac rehabilitation (CR) is a promising tool for improving functional outcome in stroke survivors, however, evidence for improving emotional health is limited. We aimed to clarify the effects of outpatient CR following in-hospital stroke rehabilitation on health-related quality of life (HRQOL) and motor function.

**Methods:** Patients with acute ischemic stroke or transient ischemic attack discharged directly home were recruited, and 128 patients (male, 92, mean age,73.5 years) who fulfilled criteria for insurance coverage of CR were divided into the CR (+) group (n=46) and CR (−) group (n=82). All patients underwent in-hospital stroke rehabilitation, and within 2 months after stroke onset, patients in the CR (+) group started a 3-month outpatient CR program of supervised outpatient sessions (1-3 times/week). Changes of motor function and HRQOL assessed by the short form-36 version 2 (SF-36) from discharge to 3 months post-discharge were compared between the two groups.

**Results:** Twenty-six patients in the CR (+) group completed the program and 66 patients in the CR (−) group were followed up at a 3-month examination. Least-square mean changes in 6-minute walk distance and isometric knee extension muscle strength were significantly higher in the CR (+) group than the CR (−) group (52.6 vs. 16.3 m; 10.1 vs. 3.50 kgf/kg). Improvement of HRQOL at 3 months was not observed in the CR (+) group.

**Conclusions:** Outpatient CR followed by in-hospital stroke rehabilitation within 2 months after stroke onset improved exercise tolerance and functional strength but not HRQOL after completion of CR.

## Introduction

Advancement in the management of acute stroke, including the use of intravenous recombinant tissue-type plasminogen activators or mechanical thrombectomy, now allows discharge directly home of about half of stroke patients admitted to the hospital after stroke onset. ^1–3^ Stroke survivors discharged home face barriers to participation in regular physical activity and spend increased time in sedentary behavior,^4,5^ which worsens comorbid health conditions and leads to stroke recurrence.^6^ Furthermore, maximal oxygen uptake level is lower in stroke survivors than in age- and gender-matched healthy controls (−53%), and this uptake level may well persist for years after stroke.^7^ Because both stroke and cardiac diseases share many of the same predisposing risk factors, such as smoking, hypertension, diabetes mellitus, physical inactivity, and dyslipidemia, modification of those risk factors combined with lifestyle intervention is recommended to prevent not only stroke recurrence but also cardiovascular diseases.^8^

Cardiac rehabilitation (CR), which is a comprehensive multidisciplinary program involving structured exercise for patients with cardiovascular diseases, has been reported to improve cardiac outcome, increase functional exercise capacity, and improve perceived health-related quality of life (HRQOL) after cardiovascular disease events.^9,10^ Exercise-based CR therapies for stroke survivors have recently been shown to improve cardiovascular endurance and functional strength.^11,12^ We reported that physical activity was decreased from premorbid levels until 90 days after stroke onset in one-third of patients discharged home after acute minor stroke or transient ischemic attack (TIA) and that apathy at discharge was a significant determinant of decreased physical activity.^13^ We thought that the use of outpatient CR, the same program used for patients with cardiovascular diseases, could improve the HRQOL of stroke survivors discharged directly home following in-hospital stroke rehabilitation, thereby improving their physical activity as well as exercise capacity. However, there is only limited evidence relating to perceptions of emotional health by stroke survivors who participate in CR.^12,14^

In the present study, we aimed to clarify the efficacy of outpatient CR by evaluating HRQOL as well as motor function in stroke survivors discharged directly home who received outpatient CR and stroke survivors discharged directly home who did not. Because insurance in Japan does not cover outpatient CR for stroke without cardiac impairment, all participants in the present study met the insurance criteria for CR coverage, i.e., had cardiac conditions.

## Methods

The present study was a single-center, retrospective, observational study approved by the Ethics Committee of the National Cerebral and Cardiovascular Center (M28-063-15) and registered with the UMIN Clinical Trials Registry (ID: UMIN000024655). All patients were notified by postings on either the home page of our hospital or the bulletin board in our rehabilitation training room that the present study would be conducted.

### Participants

We enrolled 859 patients with acute ischemic stroke or TIA admitted to the National Cerebral and Cardiovascular Center within 48 h of onset from August 2019 to March 2023 who were discharged directly home with complete independence (modified Rankin Scale of ≤1). From 859 patients, 128 patients who had a reduced left ventricular ejection fraction (≤ 40%) or an elevated B-type natriuretic peptide (BNP) level (≥ 80 pg/ml), which are the required conditions for CR insurance coverage in Japan, were recruited to receive outpatient CR (Figure 1). Of those 128 patients (male 92, mean±standard deviation, 73.5±8.4 years), 46 agreed to participate in outpatient CR and were enrolled as stroke survivors with outpatient CR [CR (+) group]. The other 82 patients were enrolled as those without outpatient CR [CR (−) group].

**Figure 1:**
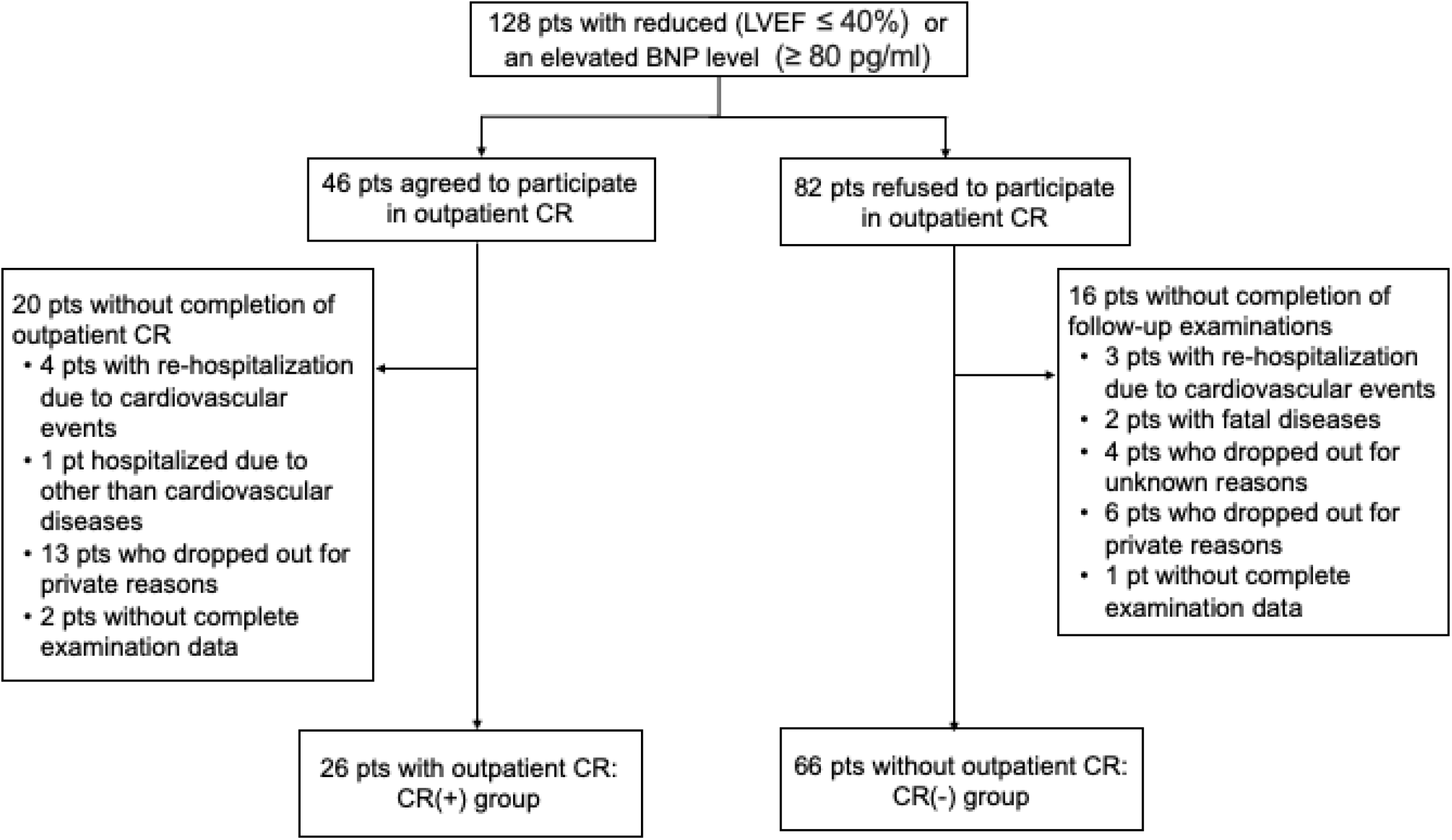
Flowchart of study participant selection. LVEF, left ventricular ejection fraction; BNP, B-type natriuretic peptide; CR, outpatient cardiac rehabilitation.

### CR program

The exercise intensity for patients with CR was determined individually to be 12–13 (“slightly difficult”) on the Borg scale of perceived exertion.^15^ All participants initiated stroke rehabilitation 2–3 days after the onset and continued it until the discharge from the stroke rehabilitation ward of our hospital. After the discharge, patients in the CR (+) group started 3-month outpatient CR with patients with cardiac diseases at the cardiac rehabilitation ward. All patients in the CR (+) group started the CR programs within 2 months after the stroke onset (mean±standard deviation, 26.3±10 days).

The outpatient CR program was the same program used to rehabilitate patients with cardiac diseases as described previously.^16^ In brief, supervised outpatient CR sessions (1–3 times/week) were provided with home exercise consisting mainly of brisk walking at a prescribed heart rate of 30-60 beats/min, 3–5 times/week, and low-intensity resistance training for 10–20 min, 2–3 times/week. Patients were encouraged to attend education classes in cardiovascular diseases, secondary prevention, and lifestyle, which were held 3–4 times each week.

### Clinical assessments

We collected baseline data, including sex, age, body mass index (BMI), the presence of hypertension, diabetes mellitus, dyslipidemia, and atrial fibrillation from medical records (Table 1). Ischemic stroke was classified into TOAST subtypes such as large-artery atherosclerosis, lacuna, cardioembolism, and other causes,^17^ and TIAs were diagnosed as previously described.^18^ Left ventricular ejection fraction (LVEF) was evaluated by echocardiography in all participants during the hospital stay. The Functional Independence Measure (FIM)^19^ for independence in the activities of daily living (with a score ranging from 18 [lowest] to 126 [highest]), Mini-Mental State Examination (MMSE) for cognitive function (with a score ranging from 0 to 30; lower scores indicating more severe cognitive dysfunction),^20^ and the Patient Health Questionnaire-9 (PHQ-9) consisting of nine questions (with a score ranging from 0 to 27; higher scores indicating the presence of more severe symptoms related to depression) as a screening tool for depression or depressive symptoms^21^ were assessed at discharge. All patients were scheduled to undergo blood tests including renal function, plasma brain natriuretic peptide (BNP), lipid profile, and glycosylated hemoglobin (HbA1c) at discharge and 3 months (3M). Renal function was assessed by the serum creatinine-based estimated glomerular filtration rate (eGFR) using equations developed for the Japanese population:^22^ eGFR (ml/min/1.73 m^2^) = 194 x (serum creatinine)^−1.094^ x (age)^−0.287^ additionally multiplied by 0.739 (for women).

**Table 1.** Patient characteristics.

|  | Total | CR (-) | CR (+) | *P value* |
| --- | --- | --- | --- | --- |
|  | (*n* =92) | (*n* = 66) | (*n* = 26) |  |
| Male | 72 (78.3) | 54 (78.3) | 20 (76.9) | 1.000 |
| Age, years | 73.4 (8.8) | 73.9 (9.0) | 72.2 (8.5) | 0.380 |
| Body Mass Index, kg/m<sup>2</sup> | 22.4 (3.7) | 22.1 (3.5) | 23.2 (4.2) | 0.237 |
| Hypertension | 65 (70.7) | 49 (71.0) | 19 (73.1) | 0.805 |
| Diabetes mellitus | 24 (26.1) | 20 (29.0) | 5 (19.2) | 0.435 |
| Dyslipidemia | 61 (66.3) | 43 (62.3) | 20 (76.9) | 0.224 |
| Atrial fibrillation | 45 (48.9) | 30 (43.5) | 17 (65.4) | 0.166 |
| Transient ischemic attack | 3 (3.3) | 2 (2.9) | 1 (3.8) | 1.000 |
| Subtypes of ischemic stroke |  |  |  | 0.586 |
| Lacuna | 17 (18.5) | 16 (23.5) | 2 (7.7) |  |
| Large-artery atherosclerosis | 10 (10.9) | 7 (10.1) | 3 (11.5) |  |
| Cardioembolism | 47 (51.1) | 33 (48.5) | 15 (57.7) |  |
| Other causes | 15 (16.3) | 11 (16.2) | 5 (19.2) |  |
| Period of hospital stay, day | 12.4 (4.2) | 11.9 (4.1) | 13.5 (4.3) | 0.117 |
| Total time of CR, min | - | 0 | 872 (450) | - |
| LVEF, % | 54.8 (8.6) | 55.8 (8.1) | 52.4 (9.5) | 0.107 |
| MMSE | 28.5 (1.7) | 28.4 (1.7) | 28.6 (1.5) | 0.688 |
| PHQ-9 | 3.9 (4.0) | 3.9 (3.9) | 3.7 (4.2) | 0.767 |
| FIM | 123 (6) | 123 (5) | 122 (7) | 0.493 |
| BNP (pg/ml) | 194 (167) | 190 (168) | 202 (168) | 0.767 |
| eGFR (ml/min/1.73 m<sup>2</sup>) | 56.3 (14.3) | 56.1 (14.6) | 56.8 (13.7) | 0.420 |
| Triglyceride (mg/dl) | 106 (71) | 110 (79) | 96 (41) | 0.275 |
| LDL cholesterol (mg/dl) | 85 (28) | 87 (28) | 79 (30) | 0.236 |
| HDL cholesterol (mg/dl) | 49 (14) | 50 (14) | 46 (14) | 0.128 |
| HbA1c (%) | 6.2 (1.1) | 6.0 (0.7) | 6.4 (1.7) | 0.273 |

Age, body mass index, LVEF, MMSE, PHQ-9, FIM, period of hospital stay, total time of CR, and blood data are mean values and standard deviations (in parentheses).
Other values are numbers and percentage (in parentheses).
CR (+), stroke survivors with outpatient cardiac rehabilitation; CR (-), stroke survivors without outpatient cardiac rehabilitation; LVEF, left ventricular ejection fraction; MMSE, Mini Mental State Examination; PHQ-9, Patient Health Questionnaire-9; FIM, Functional Independence Measure; BNP, B-type natriuretic peptide; eGFR, estimated glomerular filtration rate; LDL cholesterol, low density lipoprotein cholesterol; HDL cholesterol, high density lipoprotein cholesterol.

### Patient follow-up

Patients were followed up until 3 months after the discharge. Adverse events included: falls; symptom deterioration with NIHSS score (≥2) or cardiovascular events (stroke, TIA, myocardial infarction, unstable angina requiring revascularization, heart failure, cardiovascular death) during the exercise sessions in the outpatient CR; re-hospitalization due to cardiovascular events or surgical repair for cardiovascular diseases; death from any cause until 3 months after discharge. Adverse event data were collected until 3 months after discharge at the outpatient clinic or from medical records. Patients without any information of their conditions until 3 months after discharge were classified as having had adverse events. Recurrent stroke was assessed and recorded by experienced stroke physicians and defined as a new neurological deficit fitting the definitions for ischemic (including TIA) or hemorrhagic stroke.

### Motor function

A series of standardized, quantitative evaluations were conducted including a 6-minute walk distance according to the guidelines of the American Thoracic Society,^23^ isometric knee extension muscle strength measured with a handheld dynamometer (μTas F-1; ANIMA, Tokyo, Japan), and handgrip strength. For examination of the isometric knee extension muscle strength, patients sat in the vertical trunk position on a bench while their knee joint ankles were fixed at 90° of flexion. The dynamometer was fixed to a rigid bar. The highest strength values of the knee extensor were measured two times on each side and the averaged values on both sides were expressed relative to body weight (kgf/kg).^24^ Handgrip strength was measured on bilateral sides and mean value was obtained. These variables were examined at discharge and 3M. Changes in these 3 variables between the two time points were used as outcome measures.

### Health-related quality of life

HRQOL was assessed by the short form-36 (SF-36) version 2 questionnaire, which was translated and validated for the Japanese population,^25,26^ at discharge and the 3-month follow-up examination. The SF-36 is comprised of 36 questions that are used to measure scores in the following eight domains: physical functioning (PF), role-physical (RP), body pain (BP), social functioning (SF), general health perception (GH), vitality (VT), role-emotional (RE), mental health (MH). These eight scales are aggregated into the following three summary scores: physical component score (including positive weight for PF, RP, BP, and GH, negative weight for SF and VT), mental component score (including positive weight for MH, VT, RE, SF, and GH, negative weight for BP), and role-social component score (including positive weight for RP, RE, SF, and GH, negative weight for BP).^27^ Based on an analysis of data collected from the Japan general population survey in 2017, normative values were retrieved (mean score, 50; standard deviation, 10), with higher scores indicating better health. Changes in the three summary scores between the two time points were used as outcome measures.

### Statistical analysis

Continuous variables are presented as mean and standard deviation (SD). Categorical variables are recorded as number and percentage. We used the Student *t*-test for continuous data and Chi-square test for categorical data. The paired *t-*test was used to determine differences in each domain score and three component summary scores of the SF-36 between discharge and 3M after discharge. We used multi-regression analysis to estimate adjusted mean between-group differences in change from discharge to 3M later, and we estimated least-square means of SF-36 scores with adjustment for age, sex, BMI, and baseline SF-36 scores. We analyzed blood test data and motor function in the same way. Because participants of the present study were independent in activities of daily living, ceiling effects of measurements could not be used to show differences between before and after outpatient CR. In the analysis set for analyzing SF-36 score data, patients with the median value or less of each of the three component summary scores at discharge were included to demonstrate the effect of CR on HRQOL.

A value of p < 0.05 (2-sided) was considered to indicate a significant difference. All statistical analyses were conducted using R version 4.1.2 (R Foundation for Statistical Computing, Vienna, Austria).

## Results

In a total of 46 patients recruited as the CR (+) group, 26 patients completed the 3-month CR program (Figure 1) and were analyzed as the CR (+) group. Of the other 20 patients without completion, 4 were re-hospitalized due to cardiovascular events (2 pts. with stroke recurrence, 1 pt. with congestive heart disease, 1 pt. with surgical repair of left atrial appendage occlusion using the maze procedure), one was hospitalized and transferred to another hospital for reasons other than cardiovascular diseases, 13 dropped out due to private circumstances (for example, afraid of COVID-19 infection, caring for a family member, or going to private fitness clubs), and the remaining 2 underwent incomplete examinations. Stroke recurrence occurred in 2 patients, one before starting outpatient CR and another at home 1 month after the initiation of outpatient CR. No adverse events, such as falls, deterioration of neurological symptoms, or cardiovascular events occurred during the outpatient CR exercise sessions. Of the 82 patients without outpatient CR, 66 patients completed follow-up examinations 3M after discharge and were analyzed as stroke survivors without outpatient CR [CR (−) group]. Regarding the 16 patients without outpatient CR who did not complete follow-up, 3 patients had re-hospitalization due to cardiovascular events (2 pts. with stroke recurrence, 1 pt. with acute myocardial infarction), 2 died of cancer or COVID-19 infection, 4 dropped out for unknown reasons, 6 dropped out due to private circumstances, and the remaining one underwent incomplete examinations. Adverse events occurred in 4 of 46 patients recruited as the CR (+) group and 9 of 82 patients recruited as the CR (−) group (Figure 1). There were no significant between-group differences in the distribution of adverse events.

Regarding the 92 stroke survivors (male, 72; mean ± standard deviation, 73.4 ± 8.8 years) included in our analysis, there were no significant between-group differences in sex, age, BMI, comorbid risk factors, frequency of TIA and subtypes of ischemic stroke, and mean period of hospital stay (Table 1). The mean total CR time in 26 patients was 872 minutes (standard deviation, 450 minutes). No significant differences were observed in LVEF, MMSE, PHQ-9, and FIM at discharge. As to the blood data, there were no significant between-group differences at discharge. At 3M, BNP was decreased and HDL cholesterol increased relative to their respective levels at discharge in both groups (Suppl. Table S1).

Regarding motor function, significant increases from discharge to 3M were observed in the 6-minute walk distance and the isometric knee extension muscle strength in both groups (Figure 2). The increases in 6-minute walk distance and the isometric knee extension muscle strength were significantly larger in the CR (+) group than the CR (−) group (least-square mean change in the 6-minute walk distance, 52.6 m (95%CI: 28.4, 76.9) vs. 16.3 m [95%CI: −0.59, 33.2]; least-square mean change in isometric knee extension muscle strength, 10.1 kgf/kg [95%CI: 5.78, 14.3] vs. 3.50 kgf/kg [95%CI: 0.48, 6.51]) (Suppl. Table S2). No significant between-group difference was demonstrated in the change of handgrip strength. Component scores of HRQOL between the two time points are shown in Table 2. As an outcome measure of HRQOL, the mental component score increased significantly from discharge to 3M in the CR (−) group, but not in the CR (+) group (Figure 3, Table S3). Change of mental component score was significantly smaller in the CR (+) group than the CR (−) group (least-square mean change in the mental component score, −0.82 [95%CI: −4.42. 2.79] vs. 4.16 [95%CI: 1.68, 6.64]). There were no significant differences in either the physical component score or role-social component score between the two time points in both groups. From the further analyses of patients with median component summary scores equal to or less than 3 at discharge, the mean physical component score at 3M tended to be high compared with that at discharge, however, there were no significant differences in either the mental component score or role-social component score between the two time points in the CR (+) group (Table S4).

**Figure 2:**
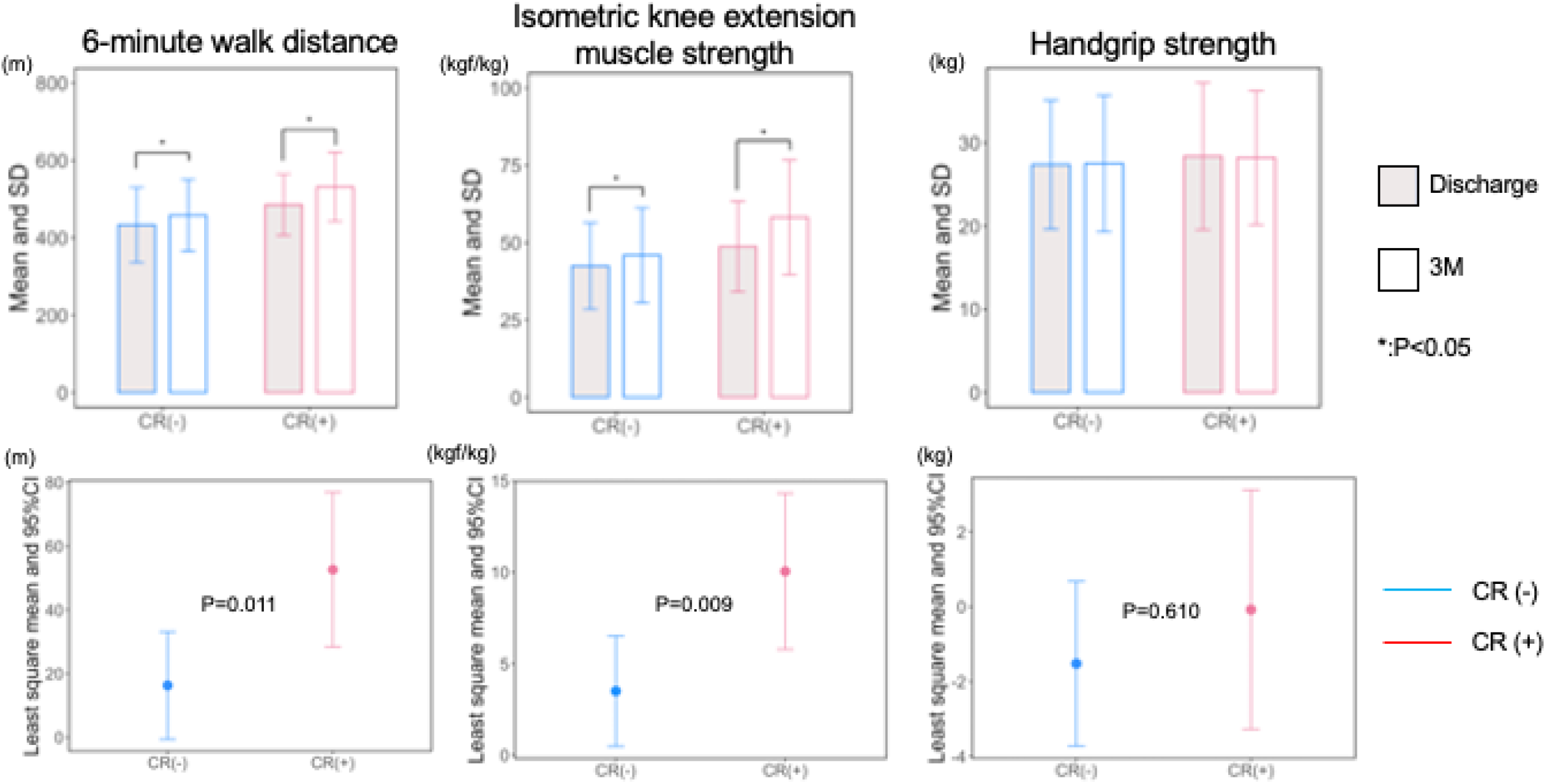
Comparison of outcome measures of motor function between CR (−) and CR (+) groups. Three bar graphs (top) show the mean ± SD of the 6-minute walk distance, isometric knee extension muscle strength, and handgrip strength, respectively, at discharge (solid bars) and at 3M (white bars). The 6-minute walk distance and isometric knee extension muscle strength were significantly higher at 3M than at discharge in both the CR (−) and CR (+) groups. Changes in these measures of motor function (bottom) between discharge and 3M expressed as least square means and 95% confidence intervals for the CR (−) group (blue line) and CR (+) group (red line). Least-square mean change in the 6-minute walk distance as well as the isometric knee extension muscle strength between the two time points in the CR (+) group is significantly higher at discharge than at 3M. CI, confidence interval; CR, outpatient cardiac rehabilitation; 3M, 3 months; *: p<0.05

**Table 2.** Comparison of SF-36 domain scores between 2 time points in each of the two groups.

|  | Mean at<br>discharge (SD) | Mean at 3M<br>(SD) | p | Least square mean *<br>(95% CI) | p |
| --- | --- | --- | --- | --- | --- |
| Physical functioning |  |  |  |  |  |
| CR (-) | 47.5 (8.2) | 47.3 (8.9) | 0.861 | -0.60 (-2.98, 1.78) | 0.806 |
| CR (+) | 48.0 (7.1) | 46.9 (8.1) | 0.425 | -1.08 (-4.52, 2.36) |  |
| Role-physical |  |  |  |  |  |
| CR (-) | 47.7 (11.9) | 44.6 (12.8) | 0.056 | -2.99 (-6.06, 0.08) | 0.292 |
| CR (+) | 46.6 (11.1) | 47.5 (9.9) | 0.690 | -0.31 (-4.79, 4.16) |  |
| Body pain |  |  |  |  |  |
| CR (-) | 50.1 (12.9) | 49.8 (11.1) | 0.847 | -0.34 (-3.22, 2.53) | 0.818 |
| CR (+) | 49.2 (12.6) | 49.6 (10.5) | 0.878 | -0.89 (-5.07, 3.30) |  |
| General health perception |  |  |  |  |  |
| CR (-) | 48.2 (8.9) | 51.5 (8.9) | 0.001 | 3.13 (0.97, 5.29) | 0.033 |
| CR (+) | 48.9 (10.6) | 48.0 (9.8) | 0.662 | -0.71 (-3.86, 2.43) |  |
| Vitality |  |  |  |  |  |
| CR (-) | 54.0 (10.1) | 56.7 (9.3) | 0.014 | 1.72 (-0.64, 4.08) | 0.071 |
| CR (+) | 55.3 (9.8) | 54.0 (11.5) | 0.548 | -1.82 (-5.26, 1.61) |  |
| Social functioning |  |  |  |  |  |
| CR (-) | 49.9 (10.9) | 52.4 (10.3) | 0.131 | 2.18 (-0.65, 5.02) | 0.31 |
| CR (+) | 48.8 (13.1) | 50.1 (9.3) | 0.648 | -0.20 (-4.34, 3.94) |  |
| Role-emotional |  |  |  |  |  |
| CR (-) | 50.2 (10.6) | 47.7 (12.3) | 0.158 | -3.03 (-6.39, 0.32) | 0.473 |
| CR (+) | 49.5 (8.8) | 45.5 (11.3) | 0.072 | -5.02 (-9.91, -0.13) |  |
| Mental health |  |  |  |  |  |
| CR (-) | 53.6 (10.1) | 57.1 (7.9) | <0.001 | 2.89 (0.98, 4.80) | 0.004 |
| CR (+) | 55.1 (9.9) | 53.1(9.8) | 0.251 | -1.79 (-4.55, 0.98) |  |

\*: Least-square mean changes from discharge (baseline) to 3M
SF-36, short form-36; 3M, 3 months; SD, standard deviation; 95%CI, 95% confidence interval
CR (-), stroke survivors without outpatient cardiac rehabilitation; CR (+), stroke survivors with outpatient cardiac rehabilitation

**Figure 3:**
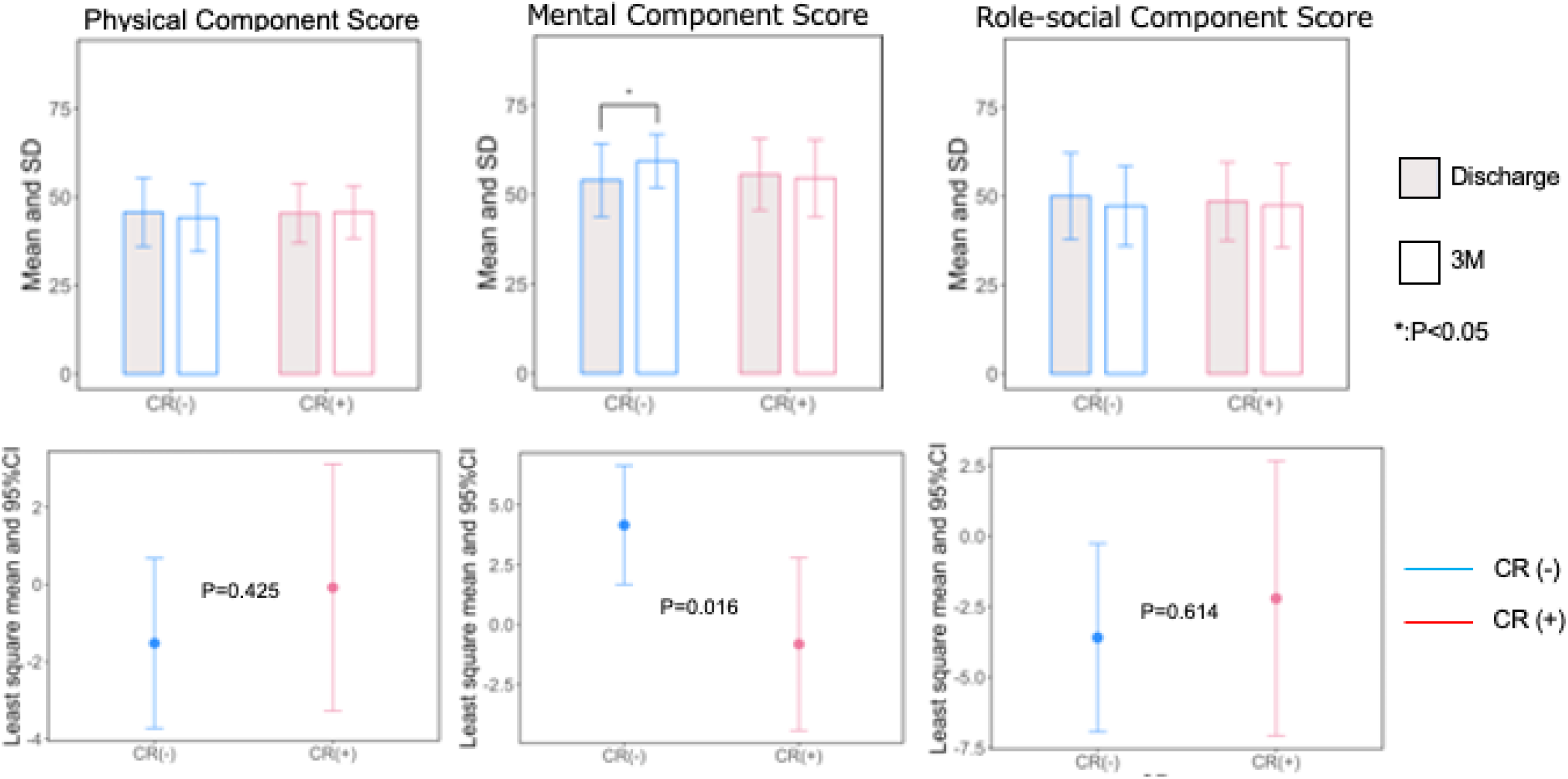
Comparison of outcome measures of health-related quality of life by the SF-36 between the CR (−) and CR (+) groups. Three bar graphs (top) show the mean ± SD of the physical component score, mental component score, and role-social component score, respectively, at discharge (solid bars) and at 3M (white bars). In the CR (−) group, the mental component score was significantly higher at 3M than at discharge. In the CR (+) group, none of the three scores changed significantly between the two time points. Changes in these health-related quality of life scores (bottom) between discharge and 3M expressed as least square means and 95% confidence intervals for the CR (−) group (blue line) and CR (+) group (red line). Least square mean change in the mental component score between the two time points is significantly lower in the CR (+) group than the CR (−) group. SF-36, short form-36; CI, confidence interval; CR, outpatient cardiac rehabilitation; 3M, 3 months; *: p<0.05

## Discussion

We showed that outpatient CR following in-hospital stroke rehabilitation improved exercise tolerance and functional strength at 3M after discharge in stroke survivors discharged directly home. However, changes of SF-36 scores between the two time points were not observed in patients participating in outpatient CR.

In the present study, mean age was 73 years old, which was about 10 years older than in the previous study,^11,12^ and no adverse events occurred during the CR exercise sessions, indicating that outpatient CR could be used for treatment of older patients with stroke. Furthermore, the timing of the initiation of CR programs after stroke onset was important in the present study. Functional recovery is known to be the greatest between 1 week and 1 month after stroke onset, still significant between 1 month and 3 months, and small between 3 and 6 months, regardless of stroke severity.^28–30^ Marzolini et al.^14^ suggested that the use of adapted CR as a standard of care practice after conventional stroke rehabilitation would be promising for a better outcome of ambulatory function in stroke survivors. We not only showed the effect of CR on motor function after stroke in survivors as previously reported,^11,12^ but also presented novel findings suggesting that the outpatient CR following conventional in-hospital stroke rehabilitation within 2 months after the stroke onset was effective in treating motor dysfunction in older stroke survivors. As for SF-36 scores, the mean physical component score was under 50 at both time points in each group. These findings indicate that the physical component score of the participants in the present study are less than the mean standardized value in the general population in Japan, which is attributable to older age and history of stroke as pointed out previously.^31,32^ Moreover, physical component score was not improved at 3M after discharge in the CR (+) group, although the 6-minute walk distance and the isometric knee extension muscle strength were significantly improved at 3M. From the additional analyses for patients with lower median physical component scores at discharge, significant improvement was also not observed in the CR (+) group. Those inconsistent findings between time points in the CR (+) group might imply discrepancies between the level of motor function restoration attained by CR and expectations for attainment as a result of participation in CR. There were also no significant differences in the mental component score and role-social component score between before and after the completion of CR and the same findings were true for patients with lower median values of each of the mental component scores and role-social component scores. Because CR is known to improve HRQOL after cardiac diseases,^9,10^ clarification is needed as to whether characteristics of stroke survivors can account for the absence of changes in HRQOL in the CR (+) group. Because patients with acute stroke participate in individual rehabilitation programs during in-hospital stroke rehabilitation in Japan, they might, after discharge, have difficulty adapting to group CR rehabilitation programs. Additionally, going to hospitals regularly for outpatient CR may be burdensome. Because participants in the present study were independent in activities of daily living and rarely had symptoms of depression according to the results of PHQ-9 screening, the effect of CR on HRQOL at 3M could hardly be detected. The small number of patients in the CR (+) group could also have contributed to this result of the present study.

Several limitations in the present study must be addressed. Given that this study had an observational single-center design, it is difficult to generalize results to a larger population. However, we showed that CR improved physical function in patients in the CR (+) group relative to those in the CR (−) group by comparison of outcome measures between patients with and without CR during the same period, but not by a one-arm before-and-after comparison study of 3-month CR programs. Second, in the present study, there was a relatively low rate of patients (35%) who agreed to participate in the outpatient CR for cardiac impairments, and a high rate of incompletion of 3-month follow-up, such as 45% in the CR (+) group and 20% in the CR (−) group. Up to one-third of participants reportedly fail to complete their CR programs.^33^ This finding could be attributable to the COVID-19 pandemic in the study period. However, there were no significant between-group differences in the distribution of adverse events until 3 months after discharge. Third, we could not evaluate psychological function (such as depression) associated with HRQOL other than by PHQ-9 examined at discharge. More studies are needed to clarify whether psychological characteristics are implicated in the changes in HRQOL by affecting participant perceptions or emotional health.

In conclusion, application of 3-month outpatient CR followed by in-hospital stroke rehabilitation within 2 months after stroke onset improved exercise tolerance and functional strength, however, no significant differences in SF-36 scores were attributable to outpatient CR completion. There may be some emotional health concerns associated with applying the outpatient CR program to stroke survivors discharged directly home. Further studies would be needed to establish strategies for encouraging stroke survivors to seek not only improvement of motor function but also emotional health. It should be also clarified whether the physical improvement shown in the present study could prevent future occurrences of cardiovascular diseases.

## Data Availability

All data are available from the corresponding author upon reasonable request.

